# Multi-omics integration predicts 17 disease incidences in the UK Biobank

**DOI:** 10.1101/2025.08.01.25332841

**Authors:** Jiawen Du, Muqing Zhou, Laura M. Raffield, Ruihai Zhou, Yun Li, Can Chen, Quan Sun

**Author notes:** Corresponding author: Quan Sun, PhD, Center for Computational and Genomic Medicine, Children’s Hospital of Philadelphia 3501 Civic Center Blvd, Colket Translational Research Building, Room 9061, Philadelphia, PA, 19104. These authors contributed equally to this work as first authors (Jiawen Du, Muqing Zhou). These authors contributed equally to this work as corresponding authors (Yun Li, Can Chen, Quan Sun). **Emails:** Jiawen Du,; Muqing Zhou,; Laura M. Raffield,; Ruihai Zhou,; Can Chen,; Yun Li,; Quan Sun.

## Abstract

**Importance:** Traditional clinical predictors for disease risks have limitations in capturing underlying disease complexity. Multi-omics technologies, such as metabolomics and proteomics, offer deeper molecular perspectives that could enhance risk prediction, but large-scale studies integrating the two omics are scarce.

**Objectives:** The primary objective is to systematically evaluate whether adding metabolomics and/or proteomics data to traditional clinical predictors improves risk prediction for 17 common incident diseases. A secondary objective is to identify key disease-related omics features.

**Data Sources and Participants:** Our study incorporated 23,776 UK Biobank participants who had complete baseline omics data for 159 NMR-based metabolites and 2,923 Olink affinity-based proteins.

**Main Outcomes and Measures:** We evaluated the model prediction of 17 incident diseases by fitting Cox proportional hazard models and obtaining Harrell’s C-index. Feature importance scores were calculated to identify key molecules contributing to each disease risk prediction.

**Results:** Adding omics data significantly improved risk prediction for all 17 diseases compared to models with clinical predictors alone (p-value < 2E-4). Proteomics-only models generally demonstrated superior predictive performance over metabolomics-only models for 14 of the 17 endpoints. We also identified key proteins, including established biomarkers like KLK3 (PSA) for prostate cancer and CRYBB2 for cataracts.

**Conclusion and Relevance:** Integration of Olink proteomics, and to a lesser extent Nightingale metabolomics, substantially improves risk prediction for a wide range of common diseases beyond established clinical factors. These findings highlight the clinical utility of proteomics for enhancing individual risk prediction and provide molecular insights into disease mechanisms, which may potentially guide future therapeutic development.

**Key Points:** *Question:* Do multi-omics profiles improve disease risk prediction compared to models using only traditional clinical risk factors and what is the best strategy to integrate metabolomics and proteomics in disease prediction?

*Findings:* In this study, we investigated 17 incident diseases across 23,776 UK Biobank individuals with complete records of both Nightingale metabolomics and Olink proteomics profiles, and found that integrating omics data significantly enhanced disease prediction over traditional approaches, with Olink proteomics consistently providing more predictive power than Nightingale metabolomics for most diseases. We also identified key proteins, including both well-established ones like KLK3 (PSA) for prostate cancer and potential novel ones like PRG3 for skin cancer. We also connected diseases with medication, socioeconomic, demographic, and lifestyle risk factors through these key proteins.

*Meaning:* Our findings suggest the potential clinical utility of integrating multi-omics in risk prediction and biomedical discoveries. To the best of our knowledge, our study is currently the largest to systematically evaluate contributions of both metabolomics and proteomics profiles to the prediction of various incident clinical endpoints.

## Introduction

Effective risk stratification is fundamental to the prevention, early detection, and management of various diseases^1^. Clinical risk assessment of disease outcome has primarily relied on established risk factors^2^, including demographic attributes, routine laboratory measurements, and sometimes lifestyle behaviors. These traditional predictors, while valuable, may not capture the underlying complex disease mechanisms. Recent advances in high-throughput omics technologies have significantly enhanced our ability to characterize biological processes at the molecular level, thereby holding the potential for improving risk assessment, disease prediction, and personalized interventions^3–5^.

Metabolomics has emerged as a robust and cost-effective method to quantify metabolites circulating in the blood. Previous studies have demonstrated the potential clinical utility of nuclear magnetic resonance (NMR)-based metabolomics, establishing their additive values beyond conventional predictors in predicting various clinical endpoints^6–9^. Similarly, proteomics, measuring proteins in various sample types including biofluids and tissues, has also been utilized to understand disease mechanisms and improve clinical risk predictions^10–12^. The UK Biobank has played a key role in exploring such disease-omics relationships at a large scale^9,10^. However, studies to comprehensively investigate the prediction power integrating these two omics remain scarce. Leveraging these complementary omics profiles holds significant promise to enhance disease prediction in clinical practice, potentially leading to more accurate diagnoses, refined prognostic prediction, and ultimately more personalized treatment strategies^13^.

To fill in the gap, we leveraged 159 metabolites (NMR based Nightingale platform) and 2,923 proteins (Affinity-based Olink 3k proteomics) measured in 23,776 individuals from the UK Biobank (UKB) to systematically evaluate whether incorporating metabolomics and proteomics data enhances prediction of 17 incident disease endpoints compared to traditional clinical predictors alone. Observing that proteomics provided robust risk prediction across most of the diseases, we further investigated top contributing proteins and identified enriched demographic, medical and socioeconomic protein correlates for each disease, leveraging external results^14^. Our study not only develops better risk prediction models, but also provides insights into a wide range of disease risk factors.

## Methods

### Omics Data and Covariates Imputation

The UKB metabolomics data, collected from the targeted Nightingale NMR based platform, were measured in 275,241 participants^9^. Proteomics profiling was conducted using Olink on blood plasma samples from 54,219 individuals^15^. We performed quality control (QC) on both omics (**Supplemental Methods**) and included 23,776 individuals as our study participants who had complete records of 159 metabolites and 2,923 proteins available (**eTables 1-2**).

We considered three nested baseline clinical predictor sets following a previous study^7^: (1) age+sex; (2) ASCVD, a set of cardiovascular-related predictors, and (3) PANEL, a more comprehensive set of clinical predictors including demographic, lifestyle, and selected laboratory measurements (**Figure 1a, eTable 3**). We assumed missing covariates were occurring at random within each recruitment center, and performed multiple imputations using chained equations (MICE)^16,17^ with random forest separately for each recruitment center (**Supplemental Methods**).

**Figure 1.**
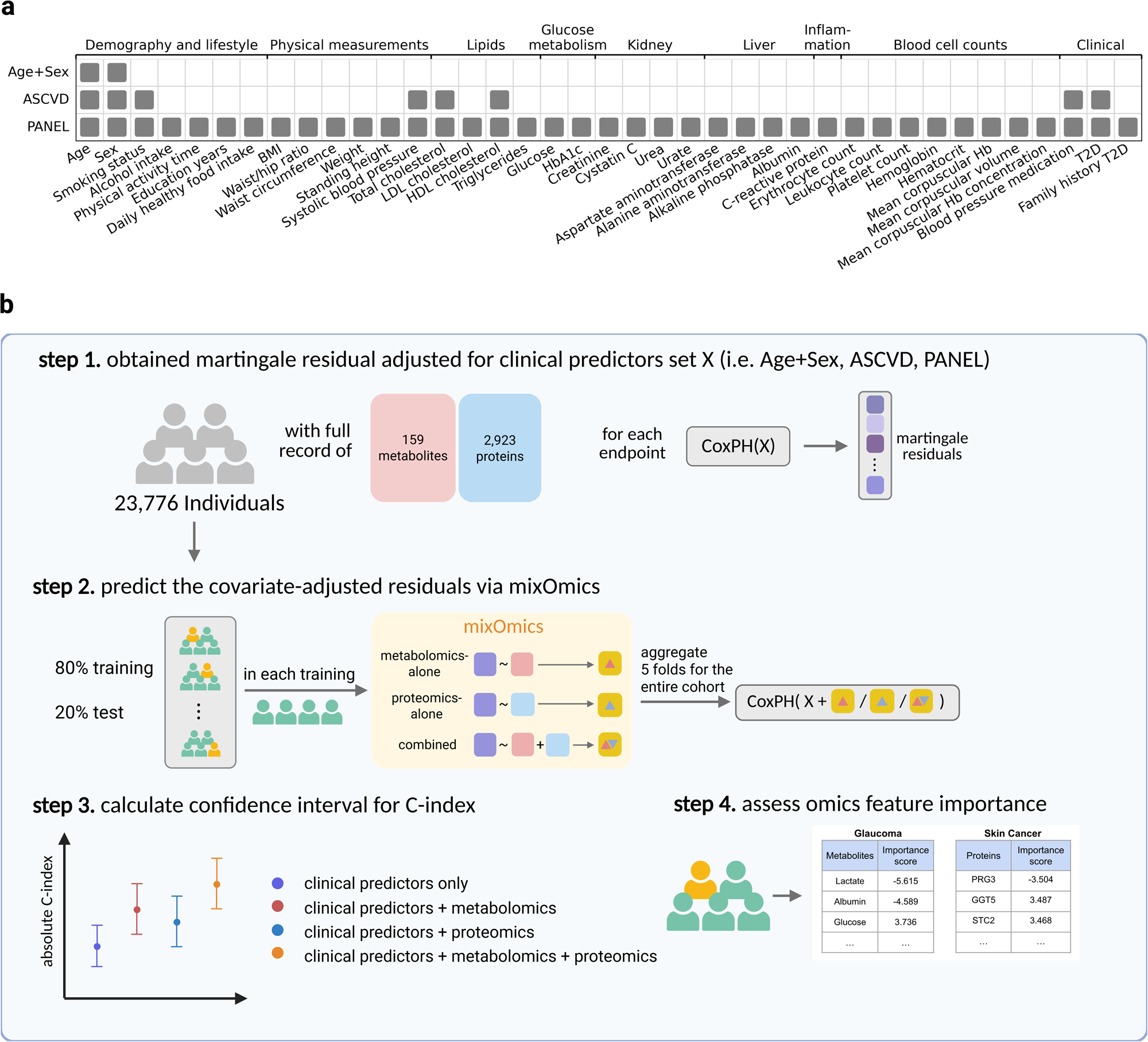
Study overview. **a.** Overview of the three baseline predictor sets. Three nested baseline clinical predictor sets were considered in this study: (1) age+sex; (2) ASCVD, a set of commonly used cardiovascular-related predictors, and (3) PANEL, a more comprehensive set of clinical predictors including demographic, lifestyle, and selected laboratory measurements. Details of (2) and (3) are included in **eTable 3**. **b.** Overview of the study design. This study included 23,776 UKB individuals with complete records of 159 metabolites and 2,923 proteins. Within this cohort, we first adjusted for one baseline clinical predictor set (X) and obtained martingale residuals using CoxPH models for each endpoint (step 1). We did both inner and outer five-fold stratified CV. For the inner CV, we split the entire cohort into five folds separately for each trait to ensure an equal number of cases in each fold, yielding the prediction across all individuals using mixOmics. Models were evaluated by adding the predicted residuals back to the CoxPH model using Harrell’s C-index (step 2). The outer CV repeated this procedure five times with different random splits to assess variations in model performance (step 3). We also identified key omics features (i.e. metabolites and proteins) for each endpoint by estimating the feature importance scores (step 4).

### Survival Endpoints Definition and Covariate Effect Adjustment

We investigated 17 endpoints, encompassing cancers, cardiovascular disorders, neurological disorders, systemic diseases, respiratory disorders, ophthalmic conditions, and musculoskeletal conditions, which were classified into 3 categories – cardiometabolic, cancers, and others (**eTable 4**). These clinical endpoints were defined by ICD10 codes following a previous publication^7^. Participants diagnosed with the specific condition prior to the baseline assessment were excluded. We confirmed that each endpoint has an incidence rate exceeding 3% in the study cohort. Analysis for highly sex-differentiated endpoints (i.e., breast cancer and prostate cancer) was restricted to the respective populations.

To control for covariates, we fitted Cox proportional hazard (PH) models for each outcome adjusting for the recruitment center and covariates. We calculated martingale residuals^18^ (**Figure 1b, step 1**), which represented the survival outcome after accounting for baseline predictor effects and were subsequently used as labels (prediction outcomes) for downstream modeling.

### Model Fitting, Evaluation, and Feature Importance Assessment

We utilized the R package mixOmics^19^ (the “block.spls” function) to perform a multivariate model training with omics data for outcome (martingale residuals) prediction. We specified 10 latent components while maintaining the default settings for the remaining parameters, and performed both inner and outer five-fold stratified cross-validation (CV). For the inner CV, we split the entire cohort into five folds separately for each trait to ensure an equal number of cases in each fold, yielding predictions across all individuals (**Figure 1b, step 2**). The outer CV repeated this procedure five times with different random splits (**Figure 1b, step 3**). We evaluated the prediction performance of three omics models, namely metabolomics-only, proteomics-only, and combined-omics, by fitting the Cox PH models again with the predicted residuals added to the model (i.e. endpoint ∼ baseline clinical predictor set + recruitment center + predicted residual). Finally, Harrell’s C-index was calculated and used as the evaluation metric (**Figure 1b, step 3**).

We also assessed important omics features contributing most to omics prediction models (**Figure 1b, step 4; Supplemental Methods**). Focusing on proteomics, we ranked all proteins by their normalized feature importance scores, separately for each endpoint and each baseline adjustment.

### Factors enriched with top contributing proteins

To further explore the potential known drivers of variance in the identified disease-associated proteins, we queried enrichment results from an external study^14^ linking proteins with multiple categories of factors, including basic demographics, medication usage, and socioeconomic factors. For each disease, we selected key proteins with an absolute normalized importance score (z-score) > 1.96 in our models, corresponding to a two-tailed p-value < 0.05, from models with the comprehensive PANEL baseline adjustment. For our enrichment queries, significance was defined as false discovery rate (FDR) < 0.05 and the corresponding enrichment odds ratio (OR) > 1.

## Results

### Characteristics of the study cohort

The final study cohort comprises 23,776 UKB participants with complete metabolomics and proteomics profiles. The mean age at enrollment was 56.9 years, ranging from 39 to 70, with 10,870 (45.7%) males. Observed incidence for the 17 investigated diseases varied, ranging from 3.0% for Dementia to 14.6% for Cataracts (**Table 1**).

**Table 1.** Characteristics of the study cohort.

| **Variable** | **Participants, No. (%) (N = 23,776)** |
| --- | --- |
| **Age, yr** |  |
| **Mean (SD)** | 56.9 (8.2) |
| **Median (range)** | 58.0 (50.0- 64.0) |
| **Sex** |  |
| **Female** | 12,906 (54.3%) |
| **Male** | 10,870 (45.7%) |
| **Body mass index** |  |
| **Mean (SD)** | 27.5 (4.8) |
| **Median (range)** | 26.8 (24.2 - 29.9) |
| **Missing** | 114 (0.5%) |
| **Smoking status** |  |
| **Current smoker** | 2,461 (10.4%) |
| **Former smoker** | 8,248 (34.7%) |
| **Never smoke** | 12,965 (54.5%) |
| **Missing** | 102 (0.4%) |
| **MACE** | 3,351 (14.09%) |
| **Heart Failure** | 1,186 (4.99%) |
| **CHD** | 3,134 (13.18%) |
| **PAD** | 1,220 (5.13%) |
| **Atrial Fibrillation** | 2,113 (8.89%) |
| **T2 Diabetes** | 2,330 (9.80%) |
| **Prostate Cancer** | 737 (3.10%) |
| **Skin Cancer** | 1,294 (5.44%) |
| **Breast Cancer** | 870 (3.66%) |
| **Dementia** | 723 (3.04%) |
| **Cataracts** | 3,475 (14.61%) |
| **Glaucoma** | 748 (3.14%) |
| **COPD** | 1,659 (6.98%) |
| **Asthma** | 2,441 (10.27%) |
| **Renal Disease** | 2,946 (12.39%) |
| **Liver Disease** | 1,628 (6.85%) |
| **Fractures** | 2,279 (9.59%) |

MACE: Major Adverse Cardiac Event; CHD: Congenital Heart Disease; PAD: Peripheral Artery Disease; COPD: Chronic Obstructive Pulmonary Disease

### Enhanced prediction accuracy including omics profiles

We first evaluated whether integrating metabolomics and/or proteomics offered improved prediction power over baseline models based solely on one of the three clinical predictor sets (**Methods**). Across all the 17 traits, adding omics data (metabolomics, proteomics, or their combination) to any baseline consistently yielded statistically significant improvement in predictive performance (**Figure 2, eTable 5**), with mean C-index increment of 0.063, 0.034 and 0.030 compared to the three baseline models, respectively (p-values ranging from 3.5E-18 to 2.1E-4). For example, compared to the baseline PANEL model, adding proteomics alone resulted in a C-index increment from 0.660 to 0.750 for prostate cancer (p-value = 1.36E-7, two-sided t-test), indicating the value of omics data beyond clinical risk factors.

**Figure 2.**
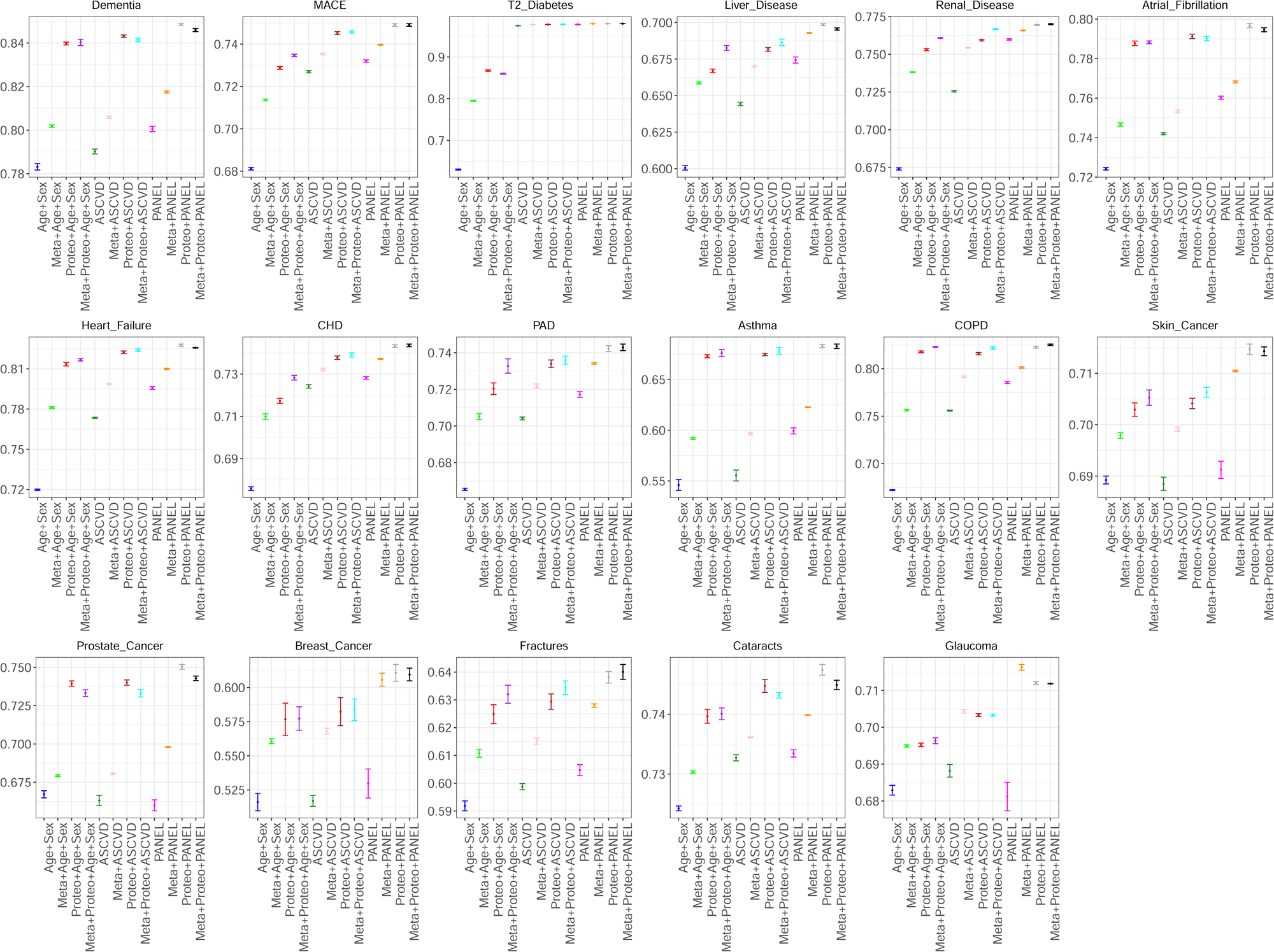
Absolute C-index for all 17 investigated endpoints. Each dot represents the mean C-index in the outer 5-fold CV with error bars indicating one standard deviation (SD). For baseline models, C-index was calculated from CoxPH models only including baseline predictors. For omics models, C-index was calculated by adding the predicted martingale residuals back to the baseline CoxPH model.

### Comparison between proteomics, metabolomics, and combined models

Comparing the individual contributions of these two omics, proteomics profiles generally showed superior predictive capability. Specifically, with PANEL baseline predictors, for 14 out of 17 traits (except for glaucoma, type 2 diabetes and breast cancer), proteomics-only models achieved significantly higher C-index values than metabolomics-only models (mean C-index difference 0.019, p-value = 1.22E-3, **Figure 2**). As an example, asthma consistently showed the greatest advantages of proteomics-only models over metabolomics-only models, with C-index differences of 0.081, 0.078 and 0.060 (p-values = 1.72E-12, 8.07E-14 and 2.90E-8) across age+sex, ASCVD and PANEL clinical predictors, respectively. Note that the number of measured proteins (n = 2,923) is substantially larger than metabolites (n = 159), which likely explains the larger contribution of proteomics models to risk predictions. Caution is needed to generalize this conclusion which may be specific to the diseases under study, and to the specific proteomics and metabolomics platforms used.

Compared to single-omics models, performance of the combined models showed variations across traits and baseline predictors, with only renal disease and chronic obstructive pulmonary disease (COPD) demonstrating consistently significant improvements across all three baseline sets (**Figure 3**). For other diseases, including both omics outperforms single-omics models for 9 of 15 traits in at least one baseline predictor set, with a trend showing less pronounced advantages of combined-omics models when including more comprehensive clinical predictors in PANEL models (**Supplemental Results, eFigures 1-2**).

**Figure 3.**
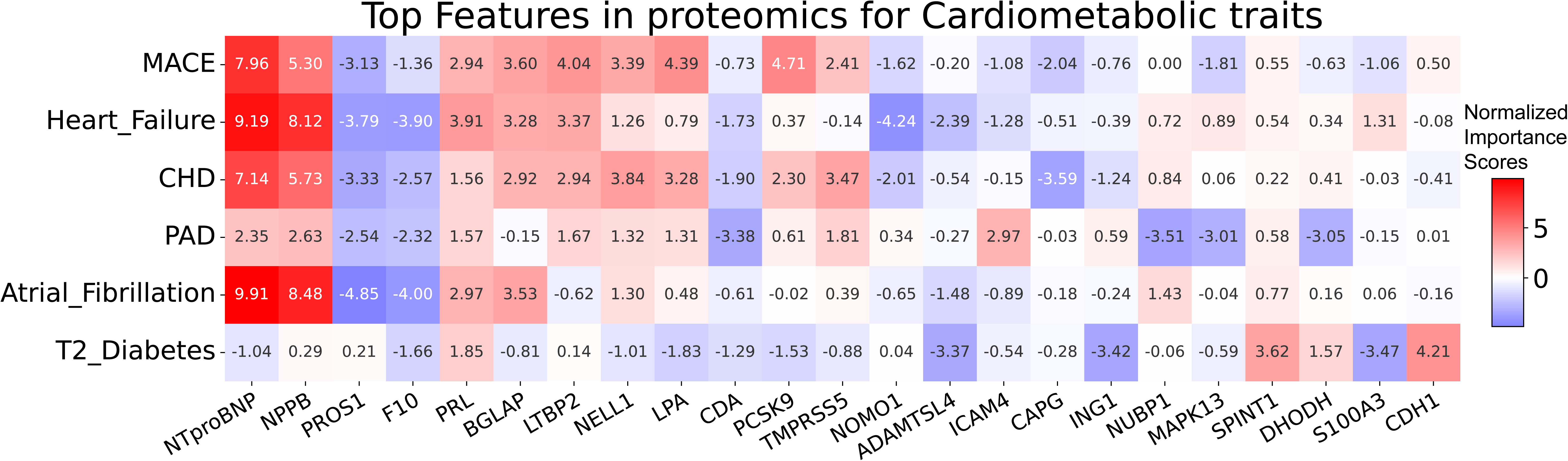
Key proteins for cardiometabolic diseases adjusting for PANEL baseline predictors. This heatmap shows identified key proteins associated with cardiometabolic disease (MACE, heart failure, CHD, PAD, AFib, and type 2 diabetes), after adjusting for the PANEL baseline clinical predictors. We selected the top 5 important proteins (regardless of direction) for each disease and displayed the union of them. Color indicates the direction of association, with intensity showing the association strength. The normalized importance scores are displayed inside each grid.

On the other hand, we only observed one trait, glaucoma, for which including metabolomics alone (C-index 0.716) showed significant while small improvements compared to either proteomics-only (C-index 0.712, p-value = 5.5E-4) or combined-omics (C-index 0.712, p-value = 2.2E-4) models with PANEL baseline predictors.

### Known disease associations confirmed by top contributing proteins and metabolites

As proteomics-only models demonstrated robust superior performance across various diseases, we then sought to identify key proteins contributing most to predictions, especially those that remained significant even after accounting for the extensive PANEL predictors (**Methods, eTable 6, eFigures 3-22**). We first highlight some notable examples with established protein-disease relationships. For example, we identified kallikrein-related peptidase 3/prostate-specific antigen (KLK3/PSA), a well-established prognostic marker, for prostate cancer^20^, which showed the strongest association among all the protein-disease pairs (z-score = 19.2, **eFigure 7**). Additionally, we confirmed a key determinant protein, Crystallin Beta B2 (CRYBB2), for cataracts^21,22^ (**eFigure 10**, z-score = 17.8); as well as Glial Fibrillary Acidic Protein (GFAP)^23^, Apolipoprotein E (APOE)^24^ and VGF^25^ for dementia (**eFigure 22**).

We also observed a consistent pattern of top-ranking proteins across cardiovascular traits (**Figure 3**). Even after adjusting for PANEL baseline, several well-established biomarkers were found prominent across atrial fibrillation (AFib), heart failure, coronary heart disease (CHD) and major adverse cardiac events (MACE). For example, brain natriuretic peptide (BNP) and N-terminal pro-brain natriuretic peptide (NT-proBNP), widely used as indicators for clinical diagnosis of cardiac dysfunction^26–29^, showed strong positive associations with all the four cardiovascular conditions (**Figure 3, eTable 6**). Moreover, a significant negative association with Protein S (PROS1) was also consistently observed, aligning with previous studies linking PROS1 deficiency to increased risks of CHD^30,31^.

At a more granular level, within these four cardiometabolic traits, we observed sub-patterns. MACE and CHD tend to have more similar proteomic signatures, sharing a significant positive association with lipoprotein(a) and proprotein convertase subtilisin/kexin type 9 (PCSK9)^32^. This sub-pattern was further confirmed when calculating correlations of protein importance scores for all proteins across traits (**Figure 4**), where we observed that cardiovascular traits formed a cluster with high correlations among each other, particularly for MACE and CHD (correlation = 0.606). Likewise, heart failure and AFib shared Coagulation Factor X (F10) which is less significant in MACE or CHD. PAD has the most distinct patterns among these conditions. While it shared the strong positive associations with BNP and NT-proBNP, PAD was also uniquely characterized by strong negative associations with cytidine deaminase (CDA), NUBP1, and MAPK13, and a positive association with ICAM4. Although these are not yet well-established biomarkers for PAD, emerging evidence suggests a plausible role for the ICAM family in the inflammatory processes that increase PAD risk^33,34^, highlighting potential avenues for further investigation. These observations were largely consistent using age+sex and ASCVD baseline sets (**eFigure 3 and 4, eTable 6**). Several other previously reported protein-disease links are summarized in **Supplemental Results**.

**Figure 4.**
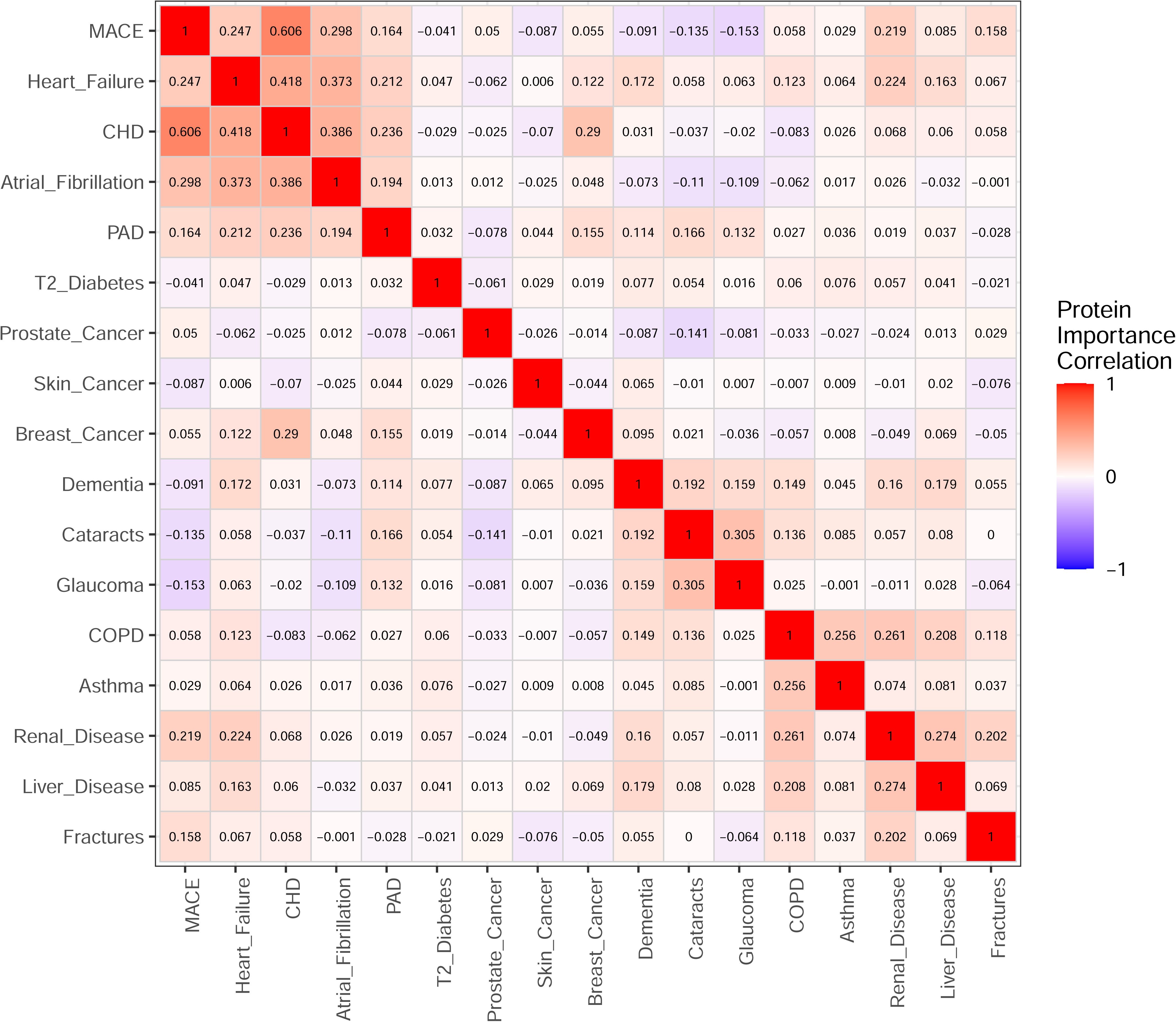
Correlations of protein importance scores for all proteins across 17 investigated endpoints. This correlation heatmap shows Pearson correlations of protein importance scores for all involved proteins across each endpoint. There are some obvious clusters across traits. For example, cardiovascular traits (i.e. MACE, heart failure, CHD, atrial fibrillation and PAD) formed a cluster showing high protein importance correlations, with the strongest relationship observed between MACE and CHD (correlation = 0.606). Other notable sub-patterns include a significant correlation between the two ophthalmologic conditions, cataracts and glaucoma (correlation = 0.305), and between the two respiratory diseases, COPD and asthma (correlation = 0.256). Similarly, liver disease and renal disease, two conditions affecting major metabolic and excretory organs, demonstrate correlation in their protein importance scores (correlation = 0.274).

For glaucoma, the only trait that metabolomics-only model performed the best, we identified lactate as the top metabolite showing a protective effect after adjusting for the PANEL predictors (**eTable 7**), which has been confirmed by previous studies^35,36^.

### Novel protein-disease relationships from top proteins

Besides well-established protein evidence, our analysis also uncovered several novel relationships in models adjusting for PANEL predictors. For example, we identified two related novel proteins for skin cancer, Proteoglycan 3 (PRG3) and gamma-glutamyltransferase 5 (GGT5). Further Mendelian Randomization (MR) analysis (**Supplemental Methods**) revealed that there is a potential causal relationship between PRG3 protein level and skin cancer (inverse-variance weighted^37^ OR=0.91, 95% CI: 0.89-0.93, p-value = 1.04E-16), with increased protein expression leading to reduced cancer risk. Sensitivity analysis using MR-Egger^38^ to correct for potential horizontal pleiotropy also supported this relationship (MR-Egger intercept p-value = 0.07; MR-Egger OR = 0.94 (0.90-0.99), p-value = 0.019).

### Enrichment of top proteins in medication usage

Leveraging our identified top contributing proteins, we queried results from an external study of UKB proteomics^14^ and found significant enrichment for proteins associated with particular medications for 11 of the 17 investigated traits, resulting in 25 distinct drug-disease associations (**eTable 8**). Six of these involved drugs uniquely associated with a single disease, while the remaining drugs were shared across multiple traits. We confirmed some expected drug-disease associations, including the widely prescribed anticoagulant Warfarin and the loop diuretic Furosemide for treating cardiovascular diseases (MACE, CHD, and AFib) (**eTable 8**). Additionally, top proteins of COPD and asthma showed enrichment for common respiratory medications, including Salbutamol (OR = 10.24, 36.32; FDR = 3.82E-8, 3.34E-26), and the combination of Salmeterol and Fluticasone Propionate (OR = 7.85, 11.7, 36.32; FDR = 3.93E-3, 2.05E-5), while asthma was also uniquely associated with Prednisolone (OR = 3.49, FDR = 2.17E-4). We note that there are few current studies exploring the medication correlates of Olink-measured proteins in true external data (i.e. not in UKB itself). As such resources become available, it would be interesting to further contextualize the putative drivers of our top proteins contributing to disease prediction.

We also uncovered new evidence supporting some existing but less well-established associations. For example, the lipid-lowering agents, Fenofibrate and Simvastatin, were associated with key proteins associated with breast cancer (OR=4.4, 2.78; FDR=2.21E-2, 9.57E-4). While some studies suggest that these two drugs tend to reduce the risk of breast cancer recurrence^39,40^, their mechanism remains unexplored. Furthermore, we found an association between Warfarin and top proteins for fracture risk (OR=3.16, FDR=1.04E-3), adding evidence to an early study suggesting the association between long-term Warfarin usage and increased osteoporotic fracture^41^. Future studies are needed to better understand these relationships.

### Enrichment of top proteins in Socioeconomics, lifestyle, and demographic factors

In addition to the enriched medications, we also identified several other factors associated with various diseases, especially for cardiometabolic traits which had the most associations with socioeconomic and lifestyle factors (including qualification, average total household income, leisure/social activities, and physical activity frequency) (**eTable 9**), consistent with previous findings that socioeconomic factors are a major determinant in cardiovascular disease risk^42^. Besides, a strong enrichment in average total household income was observed for COPD-associated proteins, which remained highly significant even after considering top proteins in the PANEL-adjusted model (OR=9.02, FDR=5.31E-7). This molecular-level finding is consistent with epidemiological data, which indicates a higher prevalence of COPD among adults with lower income levels^43^. We also confirmed expected relationships between diseases and basic demographic factors, including a strong association between incident COPD-associated proteins and smoking status (current, ever or never smoker) (OR=5.17, FDR=2.95E-19) (**eTable 10**), reinforcing the central role of smoking in COPD pathophysiology even after accounting for extensive clinical risk factors including smoking status, possibly due to the misreported smoking status or smoking intensity information (for example pack-years, which is not included in our PANEL predictor set) that is captured by top proteins.

## Discussion

In this study, we evaluated the incremental predictive performance of metabolomics and proteomics profiles, individually and in combination, upon three traditional clinical predictor sets for 17 incident diseases in 23,776 UKB participants. Compared to previous studies using metabolomics^7^ or top 5 proteins for risk prediction^11^, we achieved further enhanced predictive power for most diseases by including comprehensive proteomics and demonstrated that proteomics generally contributed more than metabolomics under these specific platforms. We also found that including both omics may not necessarily achieve better prediction under the same training sample sizes, suggesting potential model over-fitting. Our results were based on an multi-omics integration method, mixOmics^19^, while we also performed sensitivity analyses using a different method, MOGONET^44^, which led to results in highly similar patterns (**Supplemental Methods, Supplemental Results, eFigure 23**). To the best of our knowledge, our study is currently the largest to systematically evaluate contributions of both metabolomics and proteomics profiles in incident disease prediction.

We identified top contributing proteins in disease prediction models, which reinforced discoveries from previous studies focusing on proteomics-only associations and predictions^10,11^. We also uncovered some novel protein-disease associations that were to our knowledge not reported before, including a potential causal relationship of PRG3 for skin cancer. Beyond these underlying important proteins, we further identified protein-associated medication, socioeconomic and demographic factors for each disease. Our results revealed known drug-disease treatment effects, for example, the association between Warfarin and multiple disease categories covering cardiovascular, respiratory, and metabolic domains (**eTable 8**). Focusing on its pleiotropic effects, a study demonstrated that high dosage of warfarin was associated with increased expression of IL-6, COX-2, and TNF-α proteins^45^, where *IL-6* genetic variants were associated with various biomarkers and diseases (e.g, blood pressure^46^, C-reactive protein^47^, cystatin C^48^, asthma^49^, COVID-19^50^, etc.), providing a possible explanation of the multiple disease domain enrichment for warfarin-associated proteins we observed here. We also identified some less documented drug-disease relationships, suggesting potential novel repurposing drug candidates or unexpected side effects, which warrant future closer investigations.

Despite providing important insights, our study also has some limitations, most of which are due to data availability issues and point to potential future directions. First, we only included 159 metabolites measured on the NMR platform and 2,923 proteins on the Olink platform. Future studies are warranted to investigate whether our conclusions can be generalized to omics data from other platforms, for example untargeted metabolomics platforms, and to other cohorts and biobanks with differing recruitment strategies. Second, our omics profiles were collected at baseline recruitment, precluding capturing the information on disease progression. Another interesting question is to leverage longitudinal measurements, or omics trajectories, to predict disease incidence, which is not currently available. Third, we only included two omics types in this study. Future studies may integrate other omics, for example, genomics through polygenic risk scores or epigenomic markers. Fourth, we imputed the missing covariates assuming randomness within each recruitment center, which may not be true in reality. We might also have missed some cases using hospitalization records only, without incorporation of death or primary care records. Lastly, given the observation that proteomics models showed robust performance, we largely focused on top proteins for various diseases, without regard to whether top proteins were disease-shared or specific. A previous study discussed alternative strategies using disease-specific proteins (i.e., unique to each disease) to separate out proteins that have broad influences on a wide spectrum of traits^11^, which deserves further investigation of a large number of diseases at the same time. Future studies could also explore the interactive effects between metabolomics and proteomics, which is beyond the scope of this study.

In summary, we leveraged metabolomics and proteomics profiles in 23,776 UKB participants to investigate their predictive ability for 17 common incidence diseases. Our models improved risk prediction even accounting for the comprehensive PANEL clinical predictors, suggesting the values of omics profiles. Our identified key proteins and their enriched factors provide insights into potential novel disease mechanisms, repurposing drugs or unknown side effects, and a broad category of risk factors, holding promising clinical utility.

## Data Availability

UKB data are available upon request from UK Biobank (https://www.ukbiobank.ac.uk/) with approval required.

## Abbreviation used

AFib: Atrial Fibrillation
APOE: Apolipoprotein E
ASCVD: a set of cardiovascular-related predictors
BNP: brain natriuretic peptide
CDA: cytidine deaminase
CHD: Coronary Heart Disease
CI: Confidence Interval
COPD: Chronic Obstructive Pulmonary Disease
COX-2: Cyclooxygenase-2
CRYBB2: Crystallin Beta B2
CV: cross-validation
EOS: eosinophil count
FDR: false discovery rate
F10: Coagulation Factor X
GFAP: Glial Fibrillary Acidic Protein
GGT5: gamma-glutamyltransferase 5
ICAM: Intercellular Adhesion Molecule
ICD: International Classification of Diseases
IL-6: Interleukin-6
IVW: inverse-variance weighted
KLK3: kallikrein-related peptidase 3
MACE: Major Adverse Cardiac Events
MAPK13: Mitogen-activated protein kinase 13
MICE: multiple imputations using chained equations
MR: Mendelian Randomization
NMR: nuclear magnetic resonance
NT-proBNP: N-terminal pro-brain natriuretic peptide
NUBP1: Nucleotide-binding protein 1
OR: odds ratio
PAD: Peripheral Artery Disease
PANEL: a comprehensive set of clinical predictors
PCSK9: proprotein convertase subtilisin/kexin type 9
PH: proportional hazard
PRG3: Proteoglycan 3
PROS1: Protein S
PSA: prostate-specific antigen
QC: quality control
SD: standard deviation
TNF-α: Tumor necrosis factor alpha
T2 Diabetes: Type 2 Diabetes
UKB: UK Biobank

## Acknowledgments

This research has been conducted using the UK Biobank Resource under Application Number 25953. We thank the UKB participants and research team for enabling this study. This study was supported by NIH funding R01AR083790, U01HG011720, R01HL146500.

