## Supplemental Materials for "Multi-omics integration predicts 17 disease incidences in the UK Biobank"

**Table of content**

**Supplemental Methods**

**Supplemental Results**

**eFigure 1. Correlation between lipids in baseline clinical predictor sets and 159 metabolites.**

**eFigure 2. Comparison of three omics models for each clinical endpoint under three baseline clinical predictor sets.**

**eFigure 3. Top 5 key proteins for cardiometabolic traits adjusted for age_sex baseline set.**

**eFigure 4. Top 5 key proteins for cardiometabolic traits adjusted for ASCVD baseline set.**

**eFigure 5. Top 10 key proteins for cancers adjusted for age_sex baseline set.**

**eFigure 6. Top 10 Top 10 key proteins for cancers adjusted for ASCVD baseline set.**

**eFigure 7. Top 10 key proteins for cancers adjusted for PANEL baseline set.**

**eFigure 8. Top 10 key proteins for vision related traits adjusted for age_sex baseline set.**

**eFigure 9. Top 10 key proteins for vision related traits adjusted for ASCVD baseline set.**

**eFigure 10. Top 10 key proteins for vision related traits adjusted for PANEL baseline set.**

**eFigure 11. Top 10 key proteins for respiratory related traits adjusted for age_sex baseline set.**

**eFigure 12. Top 10 key proteins for respiratory related traits adjusted for ASCVD baseline set.**

**eFigure 13. Top 10 key proteins for respiratory related traits adjusted for PANEL baseline set.**

**eFigure 14. Top 10 key proteins for metabolic related traits adjusted for age_sex baseline set.**

**eFigure 15. Top 10 key proteins for metabolic related traits adjusted for ASCVD baseline set.**

**eFigure 16. Top 10 key proteins for metabolic related traits adjusted for PANEL baseline set.**

**eFigure 17. Top 15 key proteins for dementia adjusted for age_sex baseline set.**

**eFigure 18. Top 15 key proteins for dementia adjusted for ASCVD baseline set.**

**eFigure 19. Top 15 key proteins for dementia adjusted for PANEL baseline set.**

**eFigure 20. Top 15 key proteins for fractures adjusted for age_sex baseline set.**

**eFigure 21. Top 15 key proteins for fractures adjusted for ASCVD baseline set.**

**eFigure 22. Top 15 key proteins for fractures adjusted for PANEL baseline set.**

**eFigure 23. Absolute C-index for all 17 investigated traits using MOGONET**

**References**

**Supplemental Methods**

**Quality Control of the Omics Data**

For metabolomics data, we performed quality control (QC) for any metabolites flagged as QC+ in the released data from UK Biobank (UKB) by assessing the percentage below the lower limit of detection. We dropped the metabolites with such a percentage > 10%. For the remaining metabolites, we imputed missing data with the lowest observed value.

For proteomics data, we dropped proteins with missing rate greater than 20%. For the remaining proteins, we scaled the values within each batch and imputed missing values to half of the minimum observed value. Only randomized baseline participants were selected, and proteome values were subsequently inverse-normalized across batches.

**Covariates Imputation**

To handle missing covariate data, we employed Multivariate Imputation by Chained Equations (MICE), an iterative procedure that models each variable with missing data as a function of the other variables. To preserve the unique data characteristics of each recruitment location, this imputation was performed separately within each assessment center. We implemented the MICE algorithm in Python using the *scikit-learn* library, with a Random Forest estimator to capture complex, non-linear relationships. For binary covariates, imputed values were rounded to maintain their original dichotomous scale.

**Feature Importance Estimation**

To identify the most influential omics features, we employed the “block.spls” function based on Partial Least Squares (PLS) in the mixOmics^1^ package to the combined-omics model. In this model, the relationship between the predictors (i.e. omics features) and outcome is captured through a set of latent components, with loadings *L_jk_* measuring the contribution of omics feature *j* to latent component *k*. Furthermore, the Average Variance Extracted (AVE) for latent component *k* (AVE_k_) quantifies its overall explanatory power. Therefore, we defined the importance score of each feature as a weighted average of its loadings across all latent components, using the corresponding AVE values as weights. Mathematically, the importance of feature *j* can be expressed as $I_{j}=\frac{\sum_{k=1}^{K} L_{jk}*AVE_{k}}{\sum_{k=1}^{K} AVE_{k}}$ where *K* is the total number of latent components. We chose a fixed random seed 42 in sample split and treated the first fold as the test set with the remaining for training to derive feature importance scores (**Figure 1b, step d**).

**Mendelian Randomization analysis**

To investigate the potential causal relationship between PRG3 protein and the risk of skin cancer, we conducted a Mendelian randomization (MR) analysis with PRG3 protein level as exposure and skin cancer as outcome. GWAS summary statistics for PRG3 protein was collected from UK Biobank (UKB)^2^, and instrument variants were collected after performing linkage disequilibrium (LD) clumping with European TOP-LD^3^ reference panel using r^2^ > 0.001 as threshold. Summary statistics for the skin cancer were obtained from the FinnGen consortium^4^ using “Other malignant neoplasms of skin (=non-melanoma skin cancer), excluding all cancers (controls excluding all cancers)” category. After harmonizing the datasets on effect alleles and removing the SNPs that are significant in both exposure and outcome GWAS, the causal effect was primarily estimated using the random-effects inverse-variance weighted (IVW) method^5^. To assess the robustness of our results against potential violations of MR assumptions, particularly horizontal pleiotropy, we performed several sensitivity analyses using MR-Egger regression^6^.

**Sensitivity analysis using MOGONET**^7^

Besides mixOmics^1^, we also applied a deep learning method, MOGONET^7^ for sensitivity analysis. Since MOGONET was not originally designed for regression tasks with continuous labels (i.e. martingale residuals in our study setting), we slightly modified the model’s architecture to accommodate. We primarily replaced the classification-specific Cross-Entropy loss with the Mean Squared Error (MSE) loss. The terminal Softmax activation was also removed to allow for raw, continuous output values. These modifications effectively convert MOGONET from a classifier into a multi-view regression model. After this modification, we applied this model under the exact same study setting.

**Supplemental Results**

**Combined models’ performance varied by traits and baseline sets**

Compared to single-omics models, performance of the combined models showed variations across traits and baseline clinical predictors. For 11 of 17 traits (MACE, type 2 diabetes, renal disease, heart failure, CHD, PAD, COPD, skin cancer, fractures and glaucoma), the combined-omics models demonstrated significantly enhanced performance compared to both single-omics models in at least one baseline predictor set, with renal disease and COPD showing consistently significant improvements across all the three baseline sets (**Figure 3**). For example, in the age+sex adjusted model for COPD, using both omics profiles reached a C-index of 0.823, significantly outperforming metabolomics-only (C-index = 0.756, p-value = 1.03E-12) and proteomics-only (C-index = 0.818, p-value = 2.49E-5) models. For liver disease, heart failure, skin cancer and fractures, the combined-omics model showed the best performance with age+sex and ASCVD clinical predictors, but demonstrated slight inferiority to proteomics-only models with the comprehensive PANEL clinical predictors set. As the PANEL predictor set included lipids measurements, which consist of a substantial proportion of metabolomics (**eFigure 1**), the benefit of including a more comprehensive metabolite profile might be surpassed by the costs of higher degrees of freedom. A similar trend was observed that advantages of combined-omics models were less pronounced when including more comprehensive clinical predictors. Specifically, proteomics-only models significantly outperformed combined-omics models for 2 traits (type 2 diabetes and prostate cancer) with age+sex, 3 traits (dementia, prostate cancer and cataracts) with ASCVD and 6 traits (dementia, liver disease, atrial fibrillation, heart failure, prostate cancer, and cataracts) with PANEL as baseline (**eFigure 2**), although the absolute difference in C-index is minimal. For example, for the 6 traits with PANEL predictors, combined-omics models showed an average decrease in C-index of 0.002 to 0.008 compared to proteomics-only models.

**Known disease associations confirmed by top contributing proteins**

Besides cancers and cardiovascular traits, some other well-established protein-disease associations were also revealed. For the respiratory diseases including COPD and asthma, consistent patterns were observed, with Secretoglobin Family 1A Member 1 (SCGB1A1)^8,9^ and peptidoglycan recognition protein 4 (PRR4)^10,11^ identified as proteins associated with protective effects (**eFigure 13**). Metabolic traits (i.e. renal disease and liver disease) also exhibited a shared key protein pattern that remained evident even after adjustment for PANEL (**eFigure 15**). Specifically, both of them demonstrated a significant positive association with Hepatitis A Virus Cellular Receptor 1 (HAVCR1), also known as Kidney Injury Molecule 1 (KIM-1). This finding is highly consistent with the known biology of HAVCR1, as the protein is upregulated in response to acute kidney injury and hepatitis A virus infection, and its dysregulation has been observed in various liver cancers^12–14^.

**Sensitivity analysis patterns show high consistency**

The results from the sensitivity analysis using the modified MOGONET^7^ framework confirmed the substantial prediction power of omics data, as all models incorporating omics, no matter single-omics or combined, significantly outperformed the baseline model using only traditional clinical predictors (**eFigure 23**). However, within this deep learning framework, we observed minimal performance differences among proteomics, metabolomics and combined models, with slightly lower absolute values of C-indices, probably because our modified regression-based architecture was not optimized. This discrepancy highlights that computational approaches to effectively integrate multi-omics data are valuable and in pressing needs.

**Supplemental Figures**

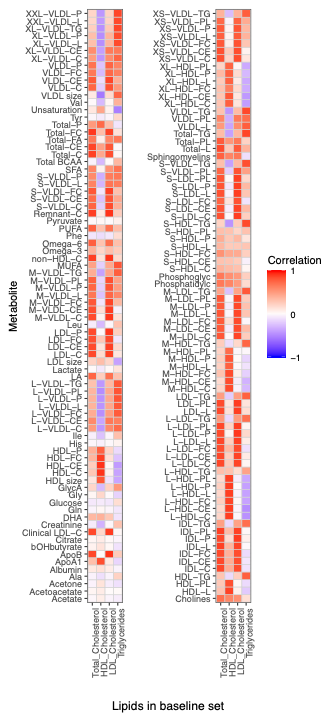

**eFigure 1.** Correlation between lipids in baseline clinical predictor sets and 159 metabolites

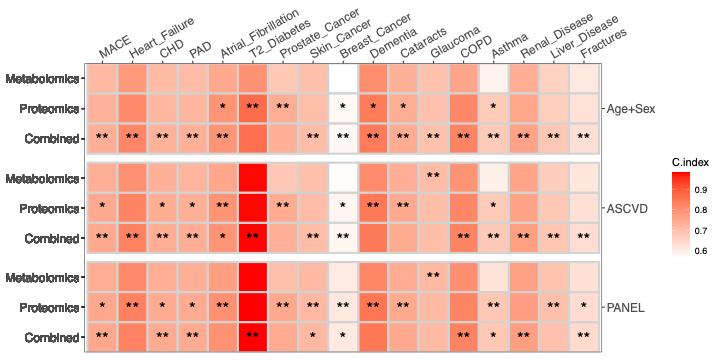

**eFigure 2.** Comparison of three omics models for each clinical endpoint under three baseline clinical predictor sets. We compared the three omics models using absolute C-index values for three baseline clinical predictor sets, including age+sex (top three rows), ASCVD (middle three rows) and PANEL (bottom three rows). Within each baseline set, the three rows correspond to the metabolomics-only, proteomics-only, and combined-omics models. Color intensity indicates the absolute C-index values. For each trait and each adjusted baseline set, two asterisks (**) indicate the best performing model across the three omics strategies (i.e. metabolomics-alone, proteomics-alone and combined), and a single asterisk (*) indicates the second best performer with no statistically significant difference from the best model.

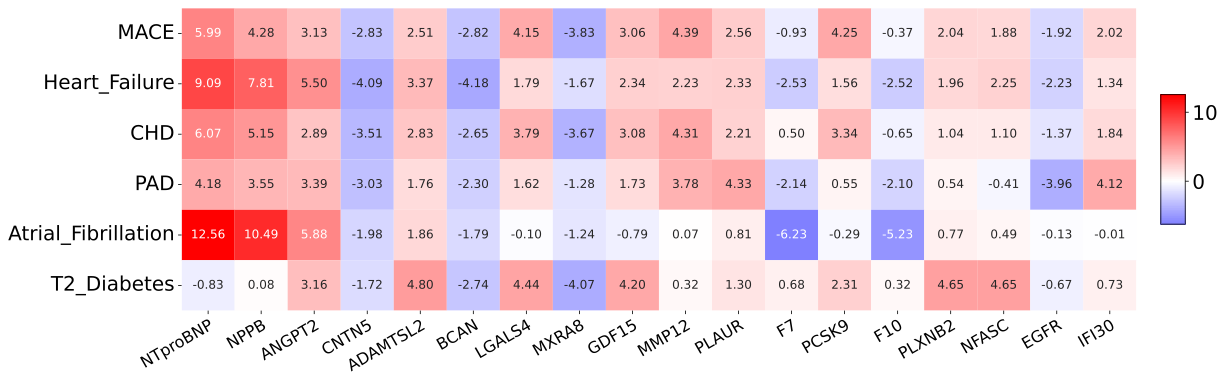

**eFigure 3.** Top 5 key proteins for cardiometabolic traits adjusted for age_sex baseline set.

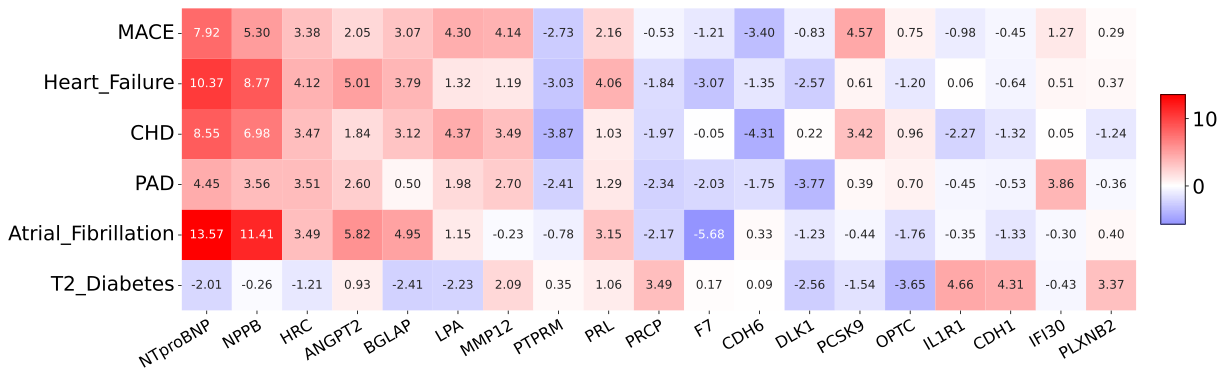

**eFigure 4.** Top 5 key proteins for cardiometabolic traits adjusted for ASCVD baseline set.

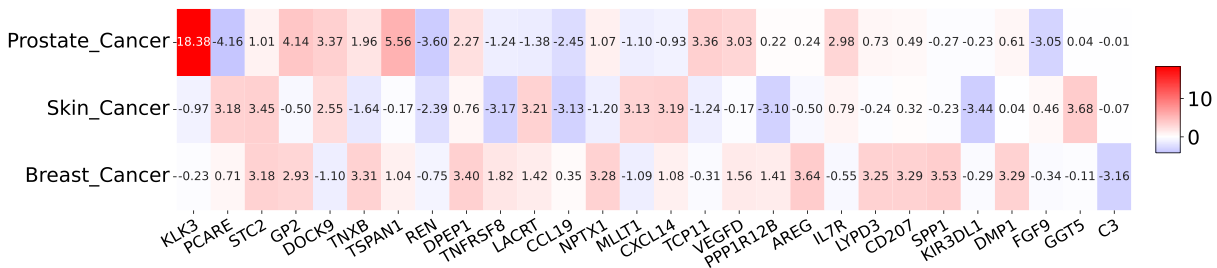

**eFigure 5.** Top 10 key proteins for cancers adjusted for age_sex baseline set.

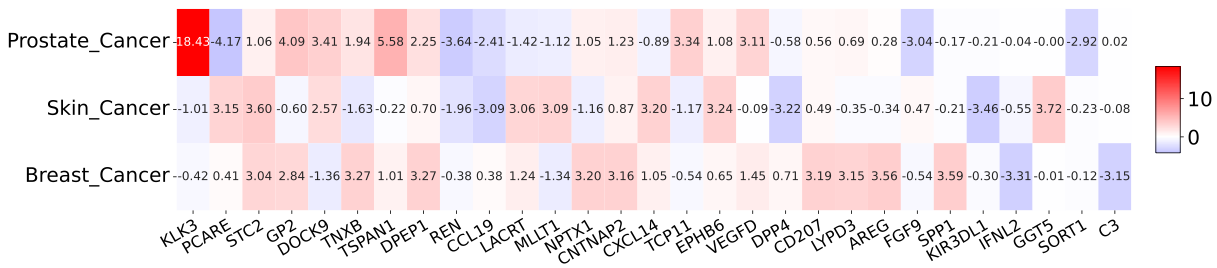

**eFigure 6.** Top 10 Top 10 key proteins for cancers adjusted for ASCVD baseline set.

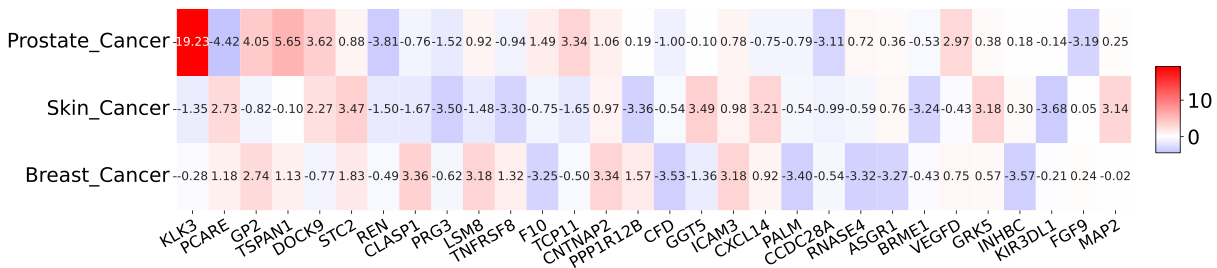

**eFigure 7.** Top 10 key proteins for cancers adjusted for PANEL baseline set.

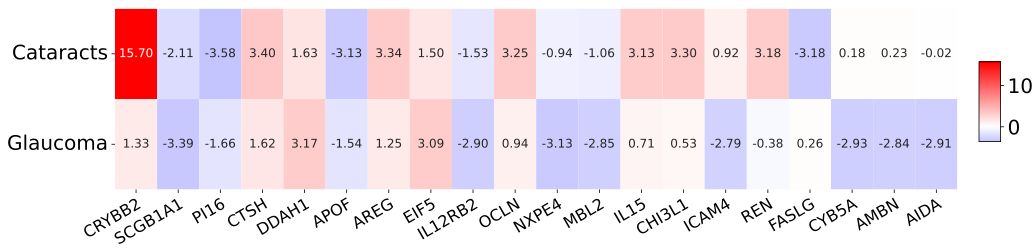

**eFigure 8.** Top 10 key proteins for vision related traits adjusted for age_sex baseline set.

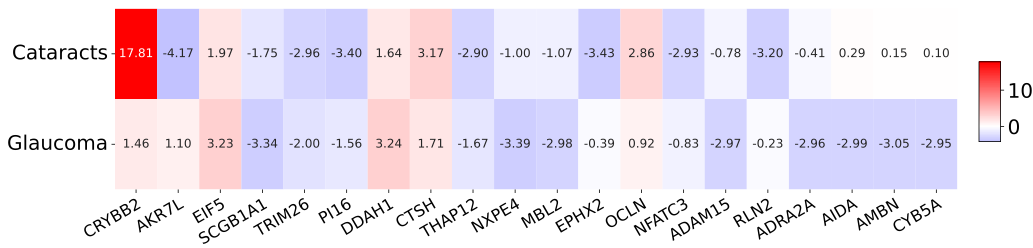

**eFigure 9.** Top 10 key proteins for vision related traits adjusted for ASCVD baseline set.

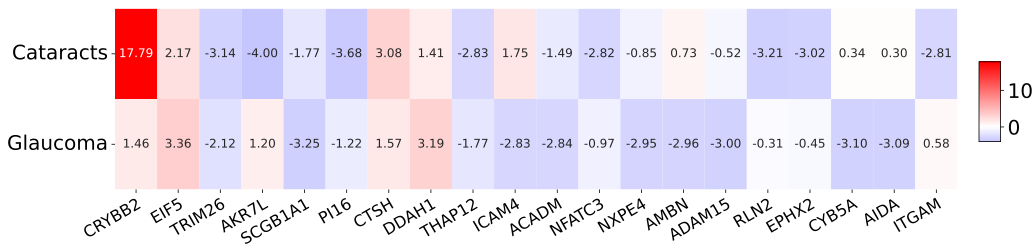

**eFigure 10.** Top 10 key proteins for vision related traits adjusted for PANEL baseline set.

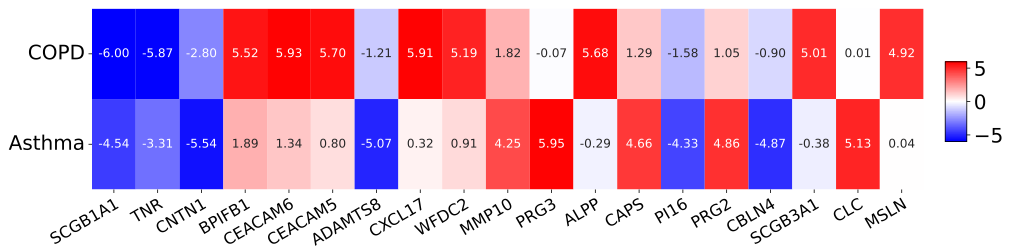

**eFigure 11.** Top 10 key proteins for respiratory related traits adjusted for age_sex baseline set.

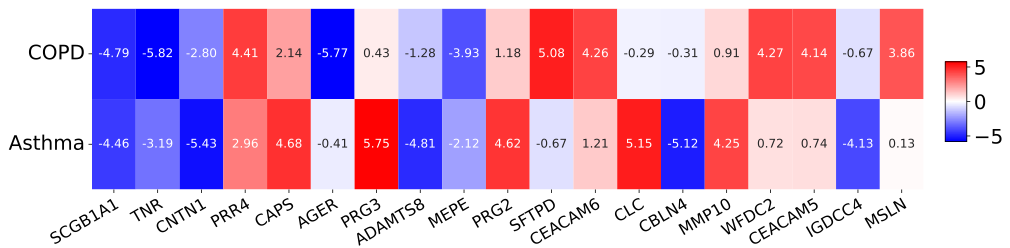

**eFigure 12.** Top 10 key proteins for respiratory related traits adjusted for ASCVD baseline set.

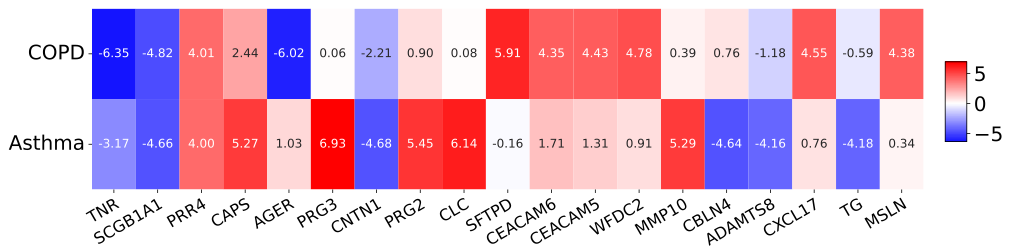

**eFigure 13.** Top 10 key proteins for respiratory related traits adjusted for PANEL baseline set.

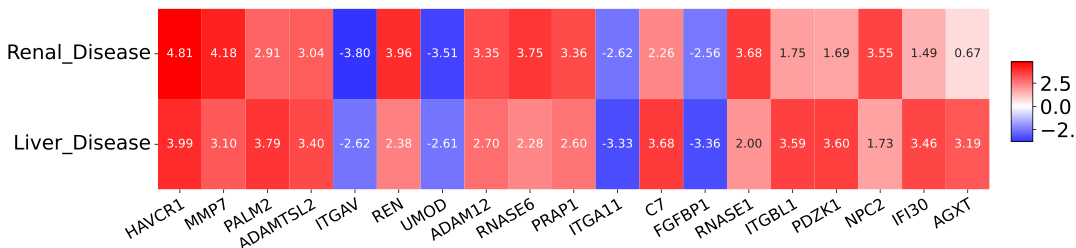

**eFigure 14.** Top 10 key proteins for metabolic related traits adjusted for age_sex baseline set.

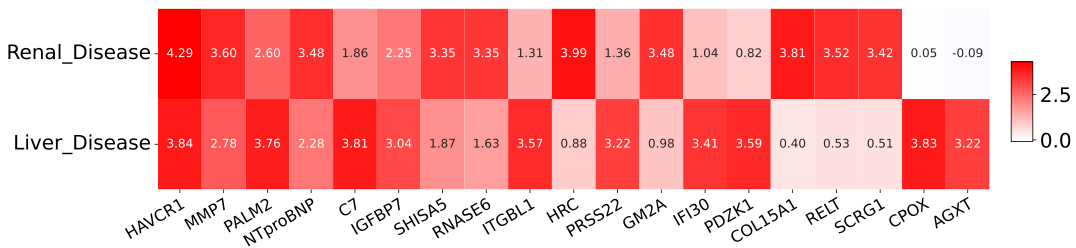

**eFigure 15.** Top 10 key proteins for metabolic related traits adjusted for ASCVD baseline set.

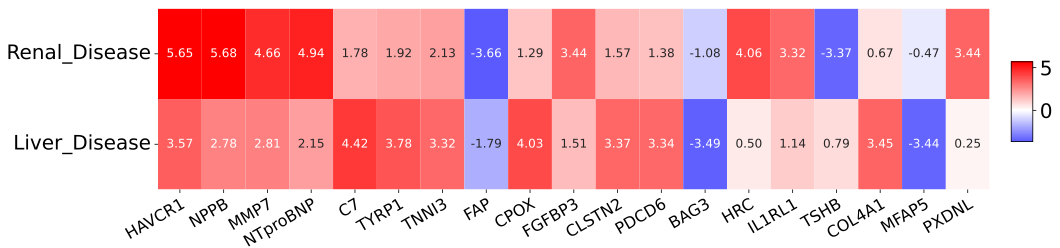

**eFigure 16.** Top 10 key proteins for metabolic related traits adjusted for PANEL baseline set.

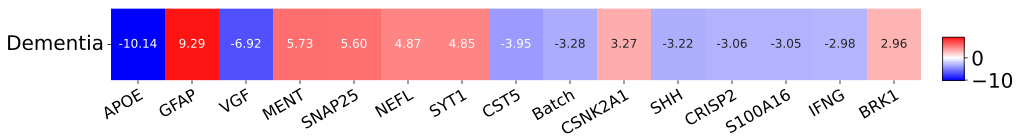

**eFigure 17.** Top 15 key proteins for dementia adjusted for age_sex baseline set.

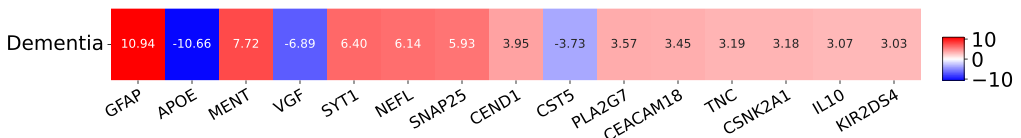

**eFigure 18.** Top 15 key proteins for dementia adjusted for ASCVD baseline set.

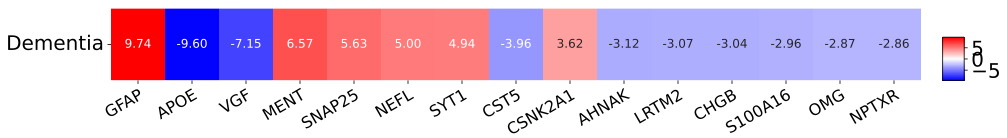

**eFigure 19.** Top 15 key proteins for dementia adjusted for PANEL baseline set.

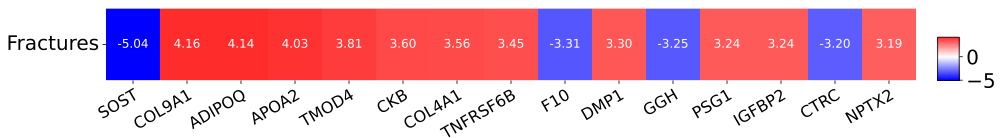

**eFigure 20.** Top 15 key proteins for fractures adjusted for age_sex baseline set.

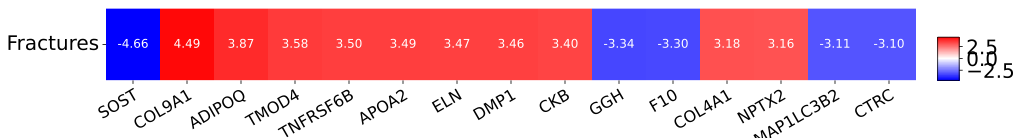

**eFigure 21.** Top 15 key proteins for fractures adjusted for ASCVD baseline set.

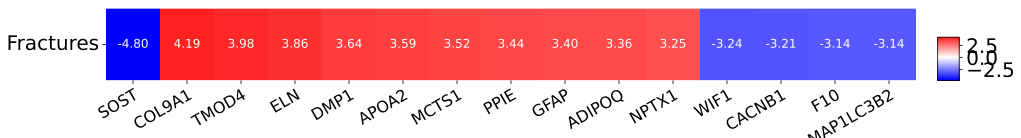

**eFigure 22.** Top 15 key proteins for fractures adjusted for PANEL baseline set.

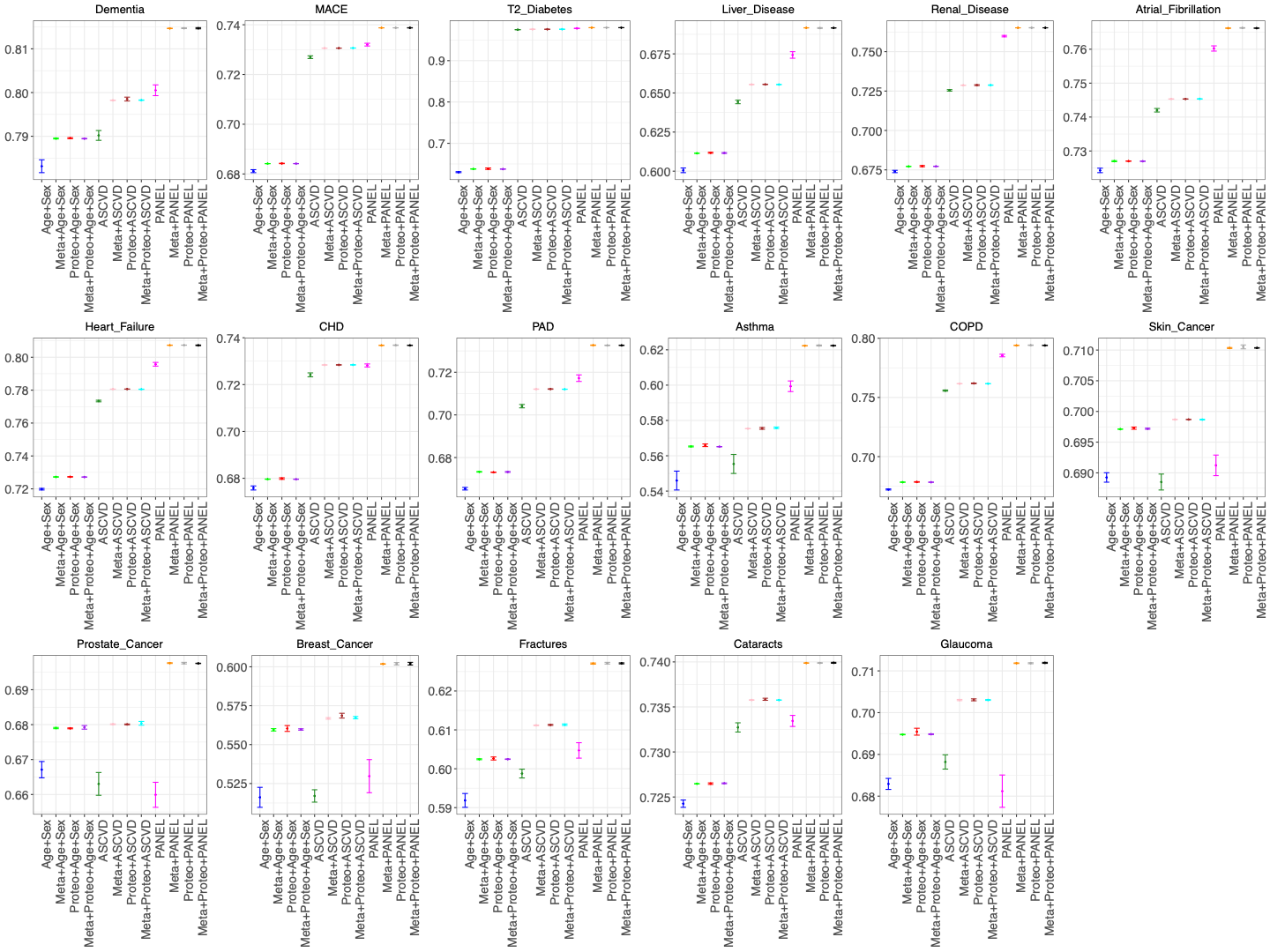

**eFigure 23. Absolute C-index for all 17 investigated traits using MOGONET**
